# Clinical characteristics and results of patients who underwent minor salivary gland biopsy with the suspicion of Sjogren syndrome

**DOI:** 10.1101/2023.01.23.23284918

**Authors:** Oznur Kutluk, Hasan Fatih Cay

## Abstract

**OBJECTIVES:** To evaluate the value and accuracy of biopsy in diagnosing Sjogren’s syndrome (SS) by evaluating the results of patients who underwent mınor salivary gland biopsy (MSGB) with the suspicion of SS.

**MATERIALS and METHODS:** The study was planned with retrospective assessment of 127 patients with biopsy performed for SS diagnosis. The clinical and laboratory characteristics of the patients who underwent MSGB were recorded from their files. SS diagnosis was placed using the rating system according to 2016 ACR/EULAR classification criteria or based on expert opinion.

**RESULTS:** 113 patients met the ınclusion criteria. 72 patients diagnosed with SS. 56 were diagnosed according to the 2016 ACR/EULAR classification criteria and 16 were diagnosed according to expert opinion. There were 57 patients with positive MSGB outcome (55 SS, 2 not SS). There were 56 patients with MSGB negative outcome (17 SS, 39 not SS). The sensitivity of MSGB for SS diagnosis was 76.4%, and specificity was 95.1%.

**CONCLUSION:** All of our patients had antibody test results, but the number of objective tests included in the classification criteria such as salivary flow rate and Schirmer was low. Although MSGB is considered an invasive technique, it appears to be a safe technique as long as it is performed by experienced professionals. It will assist in the final decision for patients with SS suspicion and who cannot undergo other objective tests.

## Introduction

Sjogren syndrome (SS) is a chronic autoimmune inflammatory disease characterized by reduced lacrimal and salivary gland functions and resulting in ocular and oral dryness. (1,2) SS may also have systemic involvement, so clinical features may be divided in two as exocrine glandular involvement and extra-glandular involvement. (3) SS is defined as primary SS when present alone, while it is called secondary SS if it accompanies other rheumatic diseases. The most common diseases associated with SS are rheumatoid arthritis (RA) and systemic lupus erythematous (SLE). (4) For diagnosis of SS, more frequently observed in women compared to men, anamnesis, Schirmer and ocular staining tests to identify dryness symptoms, salivary flow rate, serological tests and minor salivary gland biopsy (MSGB) are used. Though a variety of classification criteria were previously defined for SS, disease diagnosis may be made for clinically consistent and serologically positive people. Our aim in this study was to assess our minor salivary gland biopsy results performed with SS suspicion in our rheumatology clinic and to evaluate the place of biopsy in disease diagnosis.

## Material and method

The study was planned with retrospective assessment of 127 patients with biopsy performed for SS diagnosis in Antalya Education-Research Hospital rheumatology clinic. The files of patients with MSGB performed for SS suspicion were accessed and demographic characteristics, dryness symptoms, systemic symptoms like fatigue, arthritis and arthralgia, antibody and MSGB results were recorded.

When assessing the salivary gland biopsy performed on patients, the Chisholm and Mason classification was used to illustrate lymphocyte infiltration of the gland. (5) In minor salivary gland biopsy, lymphocyte infiltration focus was defined as 50 or above in 4 mm² and staging was based on the presence of focus.

SS diagnosis was placed using the rating system according to ACR/EULAR classification criteria published in 2016 (6) or if these criteria were not met it was based on expert opinion for patients considered to be consistent with SS clinically and serologically. The ACR/EULAR classification criteria published in 2016 are based on classifying SS in patients with at least 4 points from positive salivary gland biopsy (points (p): 3), anti-Ro antibody positivity (p: 3), ocular staining test (p: 1), Schirmer test (p: 1) and unstimulated salivary flow rate (p: 1).

## Statistical analysis and ethical aspects

Descriptive statistics are summarized as frequency (n), percentage (%), mean and standard deviation (SD) values. Assumption of normal distribution was checked with the Shapiro Wilk test. Correlations between categoric variables were assessed with the Pearson chi-square test and Fisher’s exact test. Mean ages of patients with and without diagnosis of SS were compared with the independent t test, and mean ages of diagnosis subgroups were compared with one-way ANOVA. To assess the diagnostic performance of anti-Ro, biopsy, points and biopsy+points for identification of Sjogren disease, sensitivity, specificity, negative and positive prediction values and accuracy were calculated. All analyses were performed with IBM SPSS 23.0 program (IBM Corp., Armonk, NY). P values less than 0.05 were accepted as statistically significant.

This Project was approved by the Ethics Committee in Antalya Research and Training Hospital in October 13, 2022 (protocol number 19/6).

## Results

A total of 127 patients with salivary gland biopsy were found. As the data of 14 patients could not be accessed, they were excluded from the study. Data for the remaining 113 patients were collected. When the demographic characteristics of 113 patients with MSGB performed are examined, mean age was 48.96±13.25 years, with 108 women and 5 men.

The most frequent dryness symptom was ocular dryness for 86.7%, with the most common systemic system fatigue in 80.5%. There were 34 patients with Schirmer test performed (31.1%) and 30 of these patients were positive on the Schirmer test (88.2%). Of patients, 72.6% had positive ANA test (≥1/100 titers), 74.3% were positive for anti-Ro antibody, 31.9% were positive for anti-La antibody and 33.6% were RF positive. The most frequent rheumatic disease in patients with biopsy performed was RA (n:21). Of 113 patients with MSGB performed, 72 were diagnosed with SS (63.7%). (Table 1).

**Table 1.** Demographic and clinical characteristics of the patients

| **Parameters** | **n=113 n(%)** |
| --- | --- |
| Age (years) mean±SD | **48,96±13,25** |
| Gender, n(%) |  |
| Women | **108(95,6)** |
| Men | **5(4,4)** |
| Dry mouth | 79(69,9) |
| Dry eyes | **98(86,7)** |
| Arthritis | 28(24,8) |
| Arthralgia | 82(72,6) |
| Fatigue | **91(80,5)** |
| Schimer | **34(31,1)** |
| Schimer (n=34) |  |
| Positive | **30(88,2)** |
| Negative | 4(11,8) |
| Anti-Ro |  |
| Positive | **84(74,3)** |
| Anti-La |  |
| Positive | **36(31,9)** |
| RF |  |
| Positive | **38(33,6)** |
| ANA (1/ 100, 1/320 ) | **82 (%72,6)** |
| RA | 21(18,6) |
| Focal Sialoadenitis (focus score $\geq 1$) | **57(50,4)** |
| Sjögren's syndrome | **72(63,7)** |

RF: Rheumatoid factor ANA: Antinuclear antibodies RA: Rheumatoid arthritis

The mean age of patients with SS diagnosis was 46.8 years and the mean age was lower compared to patients without diagnosis. (p<0.05) Of patients with SS diagnosis, 91.7% were positive for anti-Ro antibody, 34.7% were positive for anti-La and 26.4% of these patients were positive for both anti-Ro and anti-La antibodies. Of patients with diagnosis, anti-Ro and anti-Ro+anti-La antibody positivity were statistically significantly high, while anti-La antibody positivity alone was not significant. (Table 2) RF positivity was present in 47.2% of patients with SS diagnosis and this was significantly high compared to patients without diagnosis. Of patients, 22.2% were ANA negative, 8.2% were ANA:1/100 positive and 69.4% were ANA:1/320 positive. The most frequent ANA staining pattern was granular (68.1%). On the ANA screening test, after anti-Ro and anti-La antibodies, the most frequently observed antibody was anti-histone at 13.9% and this was significantly high in patients with diagnosis.

**Table 2.** General characteristics of patients with and without SS diagnosis

| Parameters | SS diagnosis (n=72) n(%) | Without SS (n=41) n(%) | p |
| --- | --- | --- | --- |
| Age (years) mean±SD | 46,83±12,57 | 52,71±13,74 | **0,023** |
| Anti-Ro |  |  |  |
| Positive | **66(91,7)** | 18(43,9) | **<0,001** |
| Anti-La |  |  |  |
| Positive | **25(34,7)** | 11(26,8) | 0,387 |
| Anti-Ro, Anti-La |  |  |  |
| Positive | **19(26,4)** | 1(2,4) | **0,001** |
| RF |  |  |  |
| Positive | **34(47,2)** | 4(9,8) | **<0,001** |
| ANA |  |  |  |
| Negative | 16(22,2)<sup>a</sup> | 15(36,6)<sup>a</sup> | **<0,001** |
| 1/100 | 6(8,3)<sup>a</sup> | 15(36,6)<sup>b</sup> |  |
| 1/320 | 50(69,4)<sup>a</sup> | 11(26,8)<sup>b</sup> |  |
| Granular | **49(68,1)** | 26(63,4) |  |
| RA | **16(22,2)** | 5(12,2) | 0,188 |
| Histone | 10(13,9) | 0(0) | **0,013** |
| Focal Sialoadenitis (focus score $\square \geq \square 1$) | | | |
| Negative | **17(23,6)** | 39(95,1) | **<0,001** |
| Positive | **55(76,4)** | 2(4,9) |  |

ANA: Antinuclear antibodies RA: Rheumatoid arthritis SS: Sjögren's syndrome
Independent t-test, Pearson chi-square test, Fisher's Exact test. Same letters in a row denote the lack of statistically significant difference.

Of 72 SS patients, 54 had primary SS and 18 had secondary SS (16 RA, 1 SLE and 1 scleroderma). Of patients with SS diagnosis, referrals to our clinic from other departments for research of rheumatic disease were made for 1 patient with autoimmune hepatitis, 1 with monoclonal gammopathy of undetermined significance (MGUS), 3 with leukopenia, 1 with immune thrombocytopenic purpura (ITP), 1 with optic neuritis, and 1 chronic parotitis patient. When the comorbid autoimmune diseases in patients with SS diagnosis are examined, 16 had RA, 1 had SLE, 1 had scleroderma, 2 had familial Mediterranean fever (FMF), 1 had autoimmune hepatitis, 1 had sacroiliitis, 1 had optic neuritis and 1 had MGUS.

Patients were divided into three groups, as those with 4 or more points according to the 2016 ACR-EULAR classification criteria (group 1) and patients with less than 4 points but strong suspicion of SS considering clinical and test results and diagnosed based on expert opinion (group 2) for the 72 patients with diagnosis and patients without diagnosis (group 3). (Table 3) Anti-Ro positivity was significantly high in groups 1 and 2 compared to group 3, while there was no significant difference in anti-La positivity between the groups. RF positivity was significantly high in groups 1 and 2 compared to group 3. ANA test positivity (≥1/100 titer) was significantly high in groups 1 and 2, while anti-histone positivity on the ANA screening test was higher in group 1. When the presence of arthritis patients in the groups is assessed, the presence of arthritis was significantly higher in group 2 compared to the other groups. The number of patients with MSGB positivity (focus score ≥1) was highest in group 1 and lowest in group 3. The outcomes and diagnosis distribution for patients with MSGB performed is shown in FIGURE 2. There were 57 patients with positive MSGB outcome (55 SS, 2 not SS). There were 56 patients with MSGB negative outcome (17 SS, 39 not SS). The sensitivity of MSGB for SS diagnosis was 76.4%, and specificity was 95.1%. Positive prediction value was 96.5%, negative prediction value was 69.6% and the accuracy level was 83.19%.

**Table 3.**
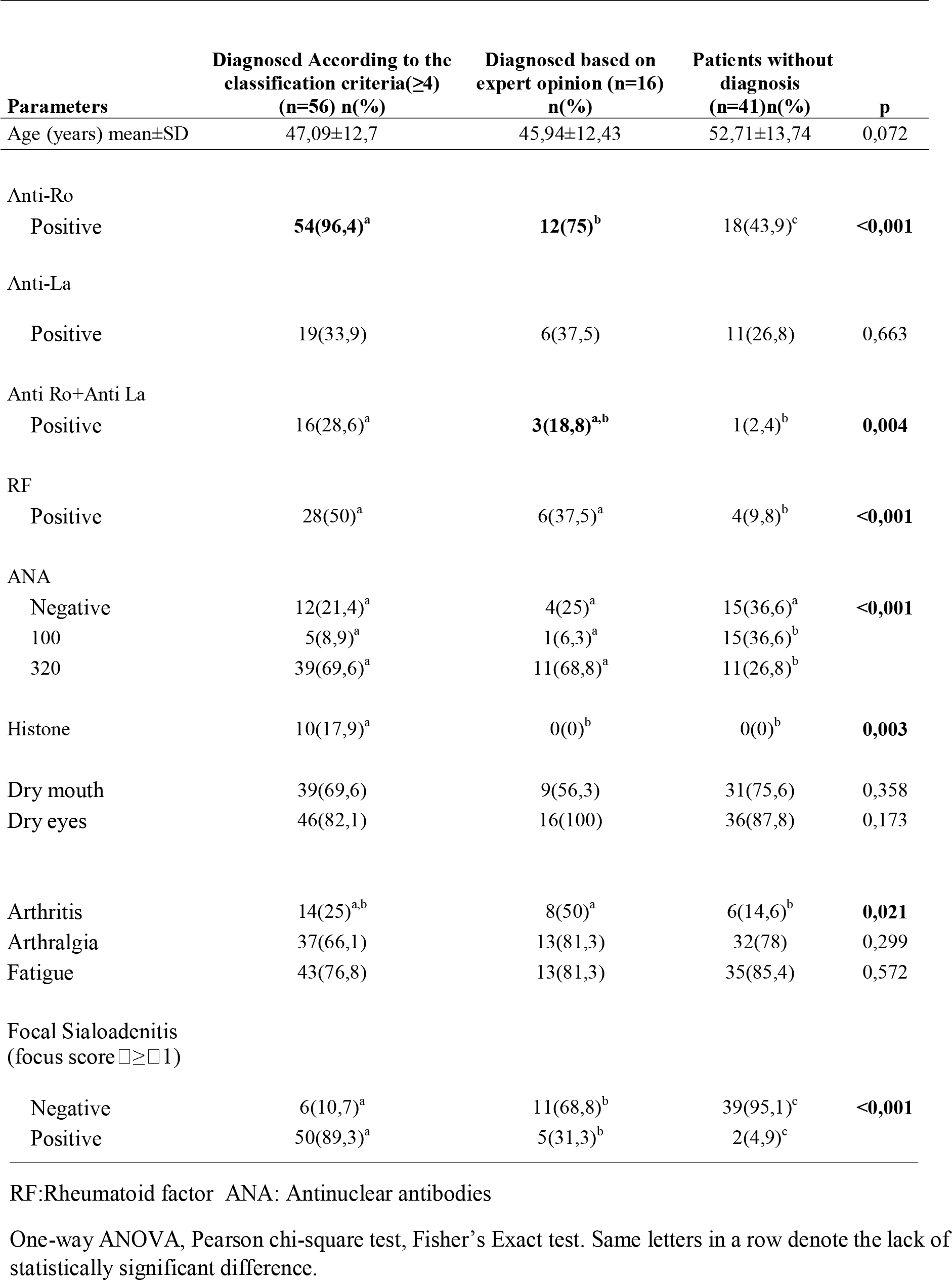
General characteristics of patients according to diagnosis subgroups

| Parameters | Diagnosed According to the<br>classification criteria($\geq 4$)<br>(n=56) n(%) | Diagnosed based on<br>expert opinion (n=16)<br>n(%) | Patients without<br>diagnosis<br>(n=41)n(%) | p |
| --- | --- | --- | --- | --- |
| Age (years) mean±SD | 47,09±12,7 | 45,94±12,43 | 52,71±13,74 | 0,072 |
| Anti-Ro |  |  |  |  |
| Positive | **54(96,4)<sup>a</sup>** | **12(75)<sup>b</sup>** | 18(43,9)<sup>c</sup> | **<0,001** |
| Anti-La |  |  |  |  |
| Positive | 19(33,9) | 6(37,5) | 11(26,8) | 0,663 |
| Anti Ro+Anti La |  |  |  |  |
| Positive | 16(28,6)<sup>a</sup> | **3(18,8)<sup>a,b</sup>** | 1(2,4)<sup>b</sup> | **0,004** |
| RF |  |  |  |  |
| Positive | 28(50)<sup>a</sup> | 6(37,5)<sup>a</sup> | 4(9,8)<sup>b</sup> | **<0,001** |
| ANA |  |  |  |  |
| Negative | 12(21,4)<sup>a</sup> | 4(25)<sup>a</sup> | 15(36,6)<sup>a</sup> | **<0,001** |
| 100 | 5(8,9)<sup>a</sup> | 1(6,3)<sup>a</sup> | 15(36,6)<sup>b</sup> |  |
| 320 | 39(69,6)<sup>a</sup> | 11(68,8)<sup>a</sup> | 11(26,8)<sup>b</sup> |  |
| Histone | 10(17,9)<sup>a</sup> | 0(0)<sup>b</sup> | 0(0)<sup>b</sup> | **0,003** |
| Dry mouth | 39(69,6) | 9(56,3) | 31(75,6) | 0,358 |
| Dry eyes | 46(82,1) | 16(100) | 36(87,8) | 0,173 |
| Arthritis | 14(25)<sup>a,b</sup> | 8(50)<sup>a</sup> | 6(14,6)<sup>b</sup> | **0,021** |
| Arthralgia | 37(66,1) | 13(81,3) | 32(78) | 0,299 |
| Fatigue | 43(76,8) | 13(81,3) | 35(85,4) | 0,572 |
| Focal Sialoadenitis<br>(focus score $\square \geq \square 1$) | | | | |
| Negative | 6(10,7)<sup>a</sup> | 11(68,8)<sup>b</sup> | 39(95,1)<sup>c</sup> | **<0,001** |
| Positive | 50(89,3)<sup>a</sup> | 5(31,3)<sup>b</sup> | 2(4,9)<sup>c</sup> |  |

RF:Rheumatoid factor ANA: Antinuclear antibodies
One-way ANOVA, Pearson chi-square test, Fisher's Exact test. Same letters in a row denote the lack of statistically significant difference.

**Figure 1.**
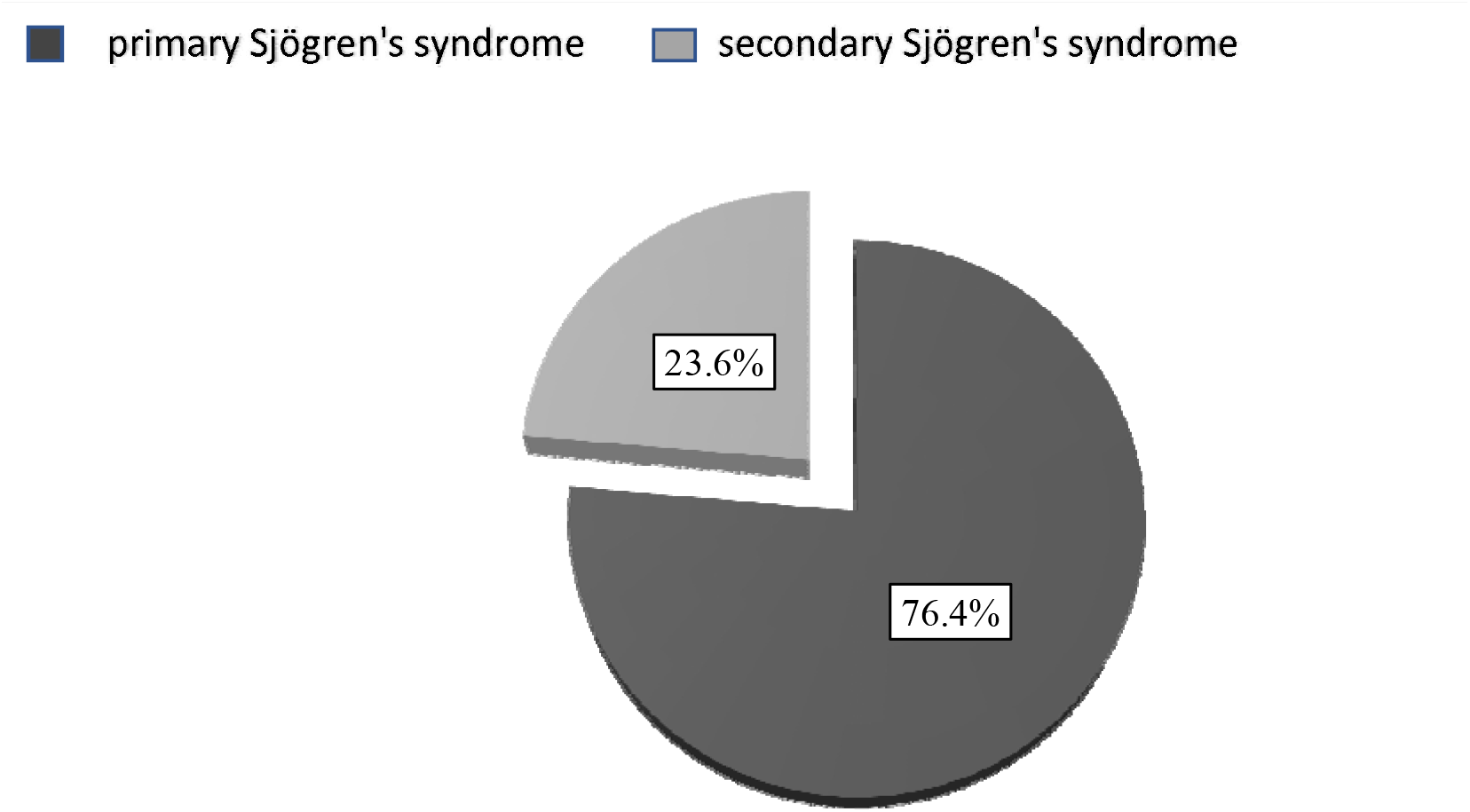
Distribution of primary and secondary Sjögren’s syndrome patients

**Figure 2:**
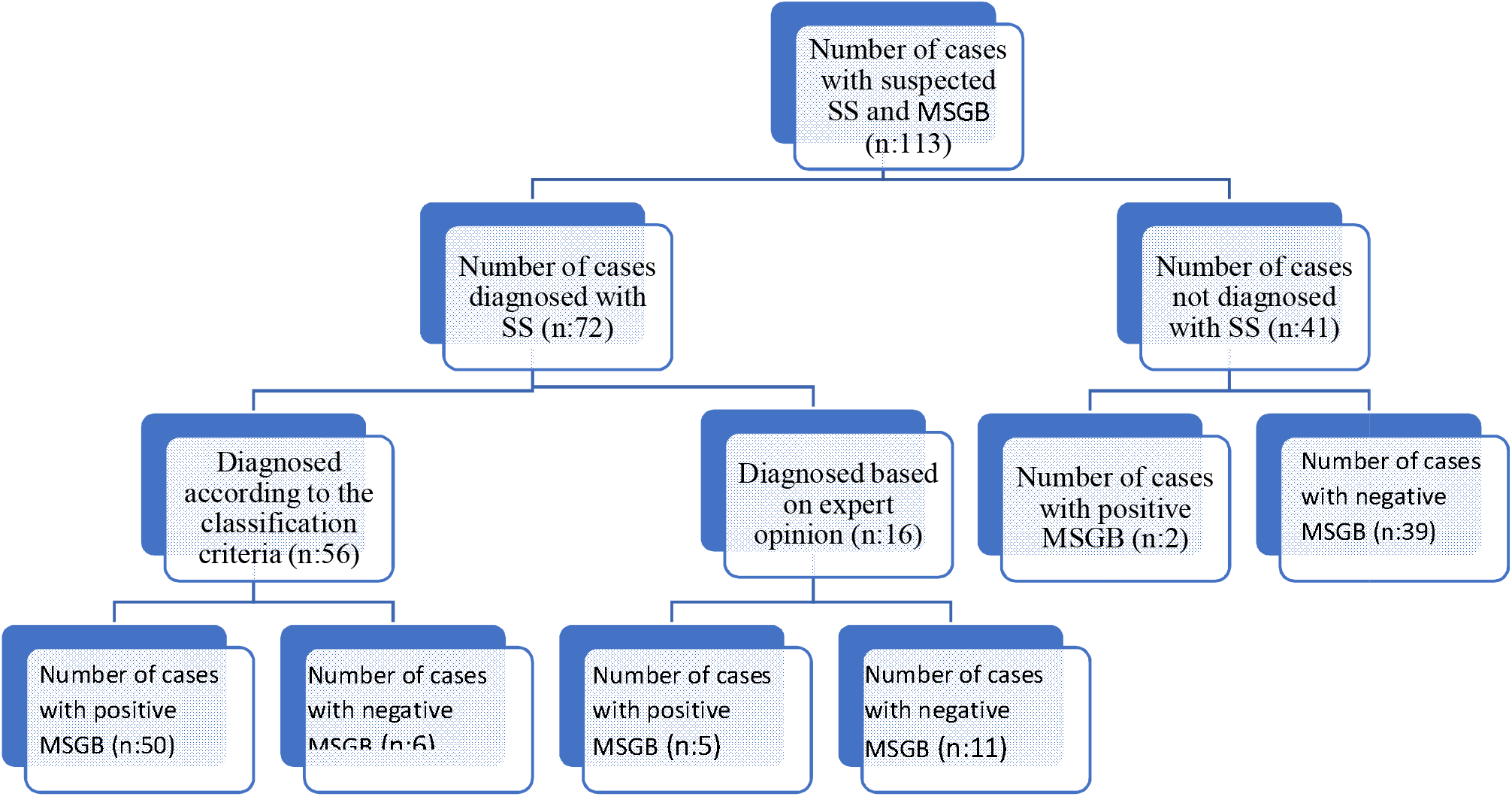
Distribution of patients who underwent MTBB by diagnosis MSGB: minor salivary gland biopsy SS:Sjögren’s syndrome

## Discussion

The aim of the study was to show the importance of MSGB performed on people with suspected SS for diagnosis of SS. In our study, SS diagnosis was made based on 2016 classification criteria or expert opinion for patients with clinical and laboratory features leading to consideration of SS but not meeting the criteria. Previous classification criteria for SS diagnosis included subjective symptoms about eye/mouth dryness from the patient and objective tests to assess dryness symptoms and antibody tests. (7) The 2016 ACR-EULAR classification criteria classify SS based on patients receiving at least 4 points on objective tests performed for patients with ocular or oral dryness. Among these tests, anti-Ro antibody and MSGB positivity are given 3 points each and the patient is classified as SS with 6 points. In our clinic, the antibody values were examined for all patients with MSGB performed for suspected SS. Though MSGB is an invasive technique, it has high points for both objective and classification criteria. Among the criteria, Schirmer test, ocular staining score and identification of reductions in unstimulated saliva flow rate are given 1 point each. These tests are performed linked to other clinical situations, are not applied in every clinic and require time. The easiest test to apply among these of the Schirmer test was only performed in 31.1% (n:34/113) of patients with biopsy due to SS suspicion, while there was no patient with ocular staining test or unstimulated saliva flow rate test performed. However, of the 72 patients with SS diagnosis, 55 were positive on MSGB. Only 20 of these 55 patients had Schirmer test performed and of these 17 had positive Schirmer test. Though the Schirmer test is easy to apply, it is notable that the number of patients undergoing this test was very low in our study.

Giovelli et al. in a study of MSGB outcomes for 216 patients with suspected SS (8) found 36.5% had positive MSGB, while in our study this value was 50.4%. ANA test positivity in the same study was 46.2%, while it was 72.6% in our study; anti-Ro positivity was 16.6%, while in our study it was much higher at 74.3%. RF positivity in the study was 18.5%, while it was 33.6% in our study. However, different to our study, this study examined the Schirmer test and unstimulated saliva flow rate. In our study, only the low rate of 31.1% of patients had Schirmer test performed, while no patient had unstimulated saliva flow rate examined. As MSGB is an invasive procedure, antibody positivity on laboratory tests performed on patients who attend with ocular/oral dryness symptoms is effective in patients with SS suspicion undergoing biopsy. In our study, the reason for higher positivity on antibody and biopsy outcomes in patients with MSGB performed may be interpreted in this way.

In our study, the number of patients with SS diagnosis was 72 (63.7%). Of these 72 patients, 54 had primary SS (76.4%) and 18 had secondary (23.6%) SS (Picture 1). The presence of SS accompanying a rheumatic disease is called secondary SS and the disease most commonly observed with secondary SS is RA. (4) For our patients, the most frequent comorbid rheumatic disease was RA (16), followed by 1 patient with SLE and 1 patient with scleroderma. A study of 50 saliva biopsies by Edelstein et al. (9) diagnosed SS in 39 patients, and of these 11 were classified as secondary SS (28% secondary SS, 72% primary SS). The most common rheumatic disease accompanying secondary SS was RA for 7 patients, similar to our study.

When the characteristics of patients with SS diagnosis are examined, anti-Ro, RF and ANA (1/320) positivity were significantly higher among patients with diagnosis compared to those without diagnosis. Though anti-La positivity was higher in the group with diagnosis, there was no statistical significance. Previous SS classification criteria (7) included anti-Ro/La, RF and ANA (1/320) positivity, while the antibodies apart from anti-Ro were removed from classification criteria in 2016. In 2016 criteria, anti-Ro positivity is assessed with 3 points, and has equal weight to MSGB within the criteria. However, as observed in our study, RF and ANA (1/320) positivity were significantly high in patients with SS diagnosis.

SS suspicion may occur not just with dryness symptoms but with extra-glandular involvement sometimes. These patients may be referred to rheumatology clinics with SS suspicion from other departments. The study by Giovelli et al. (8) found 5.5% of patients attended with symptoms like arthritis, vasculitis and polyneuropathy. In our study, this rate was 7.9% with 3 patients attending with leukopenia, 2 with multiple sclerosis, 1 with ITP, 1 with MGUS, 1 with autoimmune hepatitis and 1 with optic neuritis.

While 56 of the patients diagnosed with SS met the ACR-EULAR 2016 classification criteria (points ≥4) (group 1), 16 had SS diagnosis made based on expert opinion of clinic and serology even though they were below 4 points (group 2). All of the 16 patients in group 2 had eye dryness, while a clinical feature emerging in this group was the presence of statistically significantly more arthritis compared to the other groups. In group 2, 11 patients were MSGB negative; however they had similar rates of ANA, RF and anti-RO antibody positivity as group 1, which were higher compared to patients without diagnosis. The Schirmer test was only performed in 1 patient in group 2. The reason for these patients remaining below 4 points for the classification criteria is interpreted to be due to the lack of points due to MSGB negativity and not performing other objective tests included in the criteria (Schirmer, saliva flow rate measurement and ocular staining tests). These patients considered to have SS in terms of clinical features and serological tests were diagnosed with SS. The diagnosis and MSGB outcome distributions for patients are shown in Figure 2.

Other studies (10) examined the MSGB sensitivity and specificity and found values from 63.5-93.7% and 61.2-100%, respectively. In our study, the MSGB sensitivity was 76.4% and specificity was 95.1%, which were close to other studies.

We have no data about any complications developing after MSGB in our patients. As our study was retrospective, data related to this situation were not encountered in patient files. Complications after biopsy like pain, bruising and hemorrhage are rarely encountered; however, these symptoms generally tend to heal within a short duration.

Our study used the 2016 ACR-EULAR classification criteria for SS diagnosis; however, very few patients had the Schirmer test performed (31.1%), while the lack of any ocular staining score or unstimulated saliva flow rate test in our patient group is one of the limitations of the study.

Based on our observations, though MSGB is an invasive technique, it appears to be a safe technique as long as it is performed by experienced professionals. It will assist in the final decision for patients with SS suspicion and who cannot undergo other objective tests.

## Data Availability

All data produced in the present study are available upon reasonable request to the authors

## Consent for publication

Not applicable

## Availability of data and materials

The datasets during and/or analysed during the current study available from the corresponding author on reasonable request

## Authors’ contributions

All authors participated in all stages of preparation of the article, conception and design, acquisition of data, analysis and interpretation of data and responsible for accuracy and integrity of data. All authors read and approved the final manuscript.

